# Genetic risk and incident venous thromboembolism in middle-aged and older adults following Covid-19 vaccination

**DOI:** 10.1101/2022.04.14.22273865

**Authors:** Junqing Xie, Albert Prats-Uribe, Maria Gordillo Maranon, Victoria Y. Strauss, Dipender Gill, Daniel Prieto-Alhambra

## Abstract

**BACKGROUND:** Covid-19 vaccination has been associated with an increased risk of venous thromboembolism (VTE). However, it is unknown whether genetic predisposition to VTE is associated with an increased risk of thrombosis following vaccination.

**METHODS:** Using data from the UK Biobank, which contains in-depth genotyping data and linked vaccination and health outcomes information, we generated a polygenic risk score (PRS) using 299 genetic variants identified from a previous large genome-wide association study. We prospectively assessed associations between PRS and incident VTE after first and the second-dose vaccination separately. We conducted sensitivity analyses stratified by vaccine type (adenovirus- and mRNA-based) and using two historical unvaccinated cohorts. We estimated hazard ratios (HR) for PRS-VTE associations using Cox models.

**RESULTS:** Of 359,310 individuals receiving one dose of a Covid-19 vaccine, 160,327 (44.6%) were males, and the mean age at the vaccination date was 69.05 (standard deviation [SD] 8.04) years. After 28- and 90-days follow-up, 88 and 299 individuals developed VTE respectively, equivalent to an incidence rate of 0.88 (95% confidence interval [CI] 0.70 to 1.08) and 0.92 (95% CI 0.82 to 1.04) per 100,000 person-days. The PRS was significantly associated with a higher risk of VTE (HR per 1 SD increase in PRS, 1.41 (95% CI 1.15 to 1.73) in 28 days and 1.36 (95% CI 1.22 to 1.52) in 90 days). Similar associations were found after stratification by vaccine type, in the two-dose cohort and across the historical unvaccinated cohorts.

**CONCLUSIONS:** The genetic determinants of post-Covid-19-vaccination VTE are similar to those seen in historical data. This suggests that, at the population level, post-vaccine VTE has similar aetiology to conventional VTE. Additionally, the observed PRS-VTE associations were equivalent for adenovirus- and mRNA-based vaccines.

## Introduction

Venous thromboembolism (VTE), primarily comprising deep vein thrombosis and pulmonary embolism, is predominantly a disease of older age that affects nearly 10 million people worldwide every year and frequently leads to morbidities and death. ^1–3^ Severe acute respiratory syndrome coronavirus-2 (SARS-CoV-2) infection and coronavirus disease 2019 (Covid-19) have been recognised as novel environmental triggers for VTE. Also, a number of spontaneous thromboembolic complications were reported after Covid-19 vaccination, prompting the withdrawal of the Oxford-AstraZeneca vaccine (ChAdOx1) from several markets or restrictions on its use. Later, emerging evidence has suggested that VTE risks are substantially higher after SARS-CoV-2 infection than after vaccination, regardless of vaccine type or brand.^4^

Twins and family studies have shown that VTE is highly heritable, and a few clinical studies suggest that inherited thrombophilia can interact with various environmental risk factors, such as infectious pneumonia.^5,6^ Additionally, many genetic variants associated with VTE and their effect sizes have been identified in large-scale genome-wide association studies (GWASs), making it possible to construct a polygenic risk score (PRS) to quantify genetic predisposition to the VTE trait.

The present study aimed to assess the association between a previously validated PRS for conventional VTE and the post-Covid-19-vaccination VTE, where thrombotic events following post-Covid-19-vaccination were hypothesised to be involved in distinctive pathobiological mechanisms.

## Methods

### UK Biobank

The UK Biobank (UKBB) is a prospective cohort of over 500,000 individuals recruited from England (89%), Wales (7%) and Scotland (4%) between 2006 and 2010. Age at baseline enrolment ranged from 40 to 69 years. Comprehensive information on demographics, socioeconomics, lifestyle factors, physical metrics, and medical history were collected using a computer-based questionnaire and a standardised portfolio of measurements.^7^ Genome-wide genotyping was performed using two closely related purpose-designed arrays (the UK BiLEVE Axiom array and UK Biobank Axiom array). The genetic data have been quality controlled as described in previous studies.^8^ Over the follow-up, health-related outcomes were captured through linkage to external data sources, including primary care, hospital inpatient and death data. Additional information is available at https://www.ukbiobank.ac.uk/.

UKBB received ethical approval from the research ethics committee (National Health Service’s National Research Ethics Service North West (11/NW/0382), with all participants providing written consent. This study was conducted under Application Number 65397.

### Study population and design

For the vaccinated cohorts, all UKBB participants from England who received at least one dose of BNT162b2 or ChAdOx1Covid-19 vaccines between December 2, 2020 (i.e., vaccines approval date in the UK) and September 31, 2021 were included. Eligible participants were followed from the vaccination date (index date) to outcome, death or the end of pre-specified follow-up windows (28 and 90 days), whichever came first. The participants from Wales or Scotland were not included due to the lack of linkage to their vaccination records at the time of this analysis performed.

Two historical unvaccinated cohorts (named early-pandemic and pre-pandemic cohorts) were constructed for comparison. For the early-pandemic cohort, the observational period started from March 23 2020 (the announcement of the first national lockdown in the UK, index date) to December 1 2020 (the last day before Covid-19 vaccines approval). In contrast, the pre-pandemic cohort was followed one year earlier, from March 23 2019 (index date) to March 23 2020. In addition, a Covid-19 infection cohort was curated with the date of infection as index date. People with historical VTE at the study entry date were excluded for all study cohorts.

### Polygenic risk score

We derived polygenic risk scores (PRS) for VTE as a weighted sum of risk alleles, using summary statistics of 299 single nucleotide polymorphisms (SNPs) from a genome-wide association study (GWAS) on VTE, which included the two clinically validated mutations: factor V Leiden p.R506Q and prothrombin G20210A.^9^ Given that the selected GWAS sample included UKBB participants, we conducted a sensitivity analysis using a newly generated alternative PRS based on a meta-analysis of 12 GWASs that did not cover UKBB participants.^10^ We standardised the continuous PRS by *z*-transformation to achieve a zero mean and standard deviation of 1 based on the entire UKBB population.

Details on data manipulation and completed lists of SNPs included in the primary PRS and alternative PRS are provided in the **sTable 1** and **sTable 2**.

**Table 1:** Baseline characteristics by the genetic risk categories (one dose).

|  |  | High PRS groups |  |  |
| --- | --- | --- | --- | --- |
|  | Overall | Top 33% | Top 20% | Top 5% |
| **Number** | 359,310 | 119,770 | 71,862 | 17,965 |
| **Demographics** |  |  |  |  |
| Age, mean (sd) | 69.05 (8.04) | 69.10 (8.01) | 69.14 (8.02) | 69.11 (8.02) |
| Sex, male (%) | 160327 (44.6) | 53178 (44.4) | 31766 (44.2) | 7909 (44.0) |
| BMI, mean (sd) | 27.30 (4.69) | 27.29 (4.69) | 27.29 (4.69) | 27.26 (4.69) |
| **Socio-economic status, mean (sd)** |  |  |  |  |
| IMD (Indices of Multiple Deprivation) | 17.18 (13.62) | 17.00 (13.50) | 16.91 (13.43) | 16.71 (13.27) |
| Income score | 0.11 (0.09) | 0.11 (0.09) | 0.11 (0.09) | 0.11 (0.09) |
| Employment score | 0.09 (0.06) | 0.09 (0.06) | 0.09 (0.06) | 0.08 (0.06) |
| Health score | -0.11 (0.84) | -0.12 (0.84) | -0.13 (0.84) | -0.14 (0.84) |
| Education score | 15.23 (15.84) | 15.14 (15.75) | 15.09 (15.74) | 14.98 (15.70) |
| Housing score | 19.76 (10.12) | 19.64 (10.08) | 19.58 (10.07) | 19.47 (10.10) |
| Crime score | -0.06 (0.77) | -0.06 (0.77) | -0.07 (0.77) | -0.07 (0.78) |
| Living environmental score | 18.28 (15.07) | 18.15 (14.96) | 18.08 (14.89) | 17.93 (14.66) |

Indices of Multiple Deprivation offer a more complex and detailed view of deprivation, based on more factors than the Townsend index. All scores has been scaled to 0 to 1, 0 to 100, or even distributions standardized around 0, with higher values indicating more deprived. Details of individual score has been described in the GOV.UK (https://www.gov.uk/government/collections/english-indices-of-deprivation).

**Table 2:**
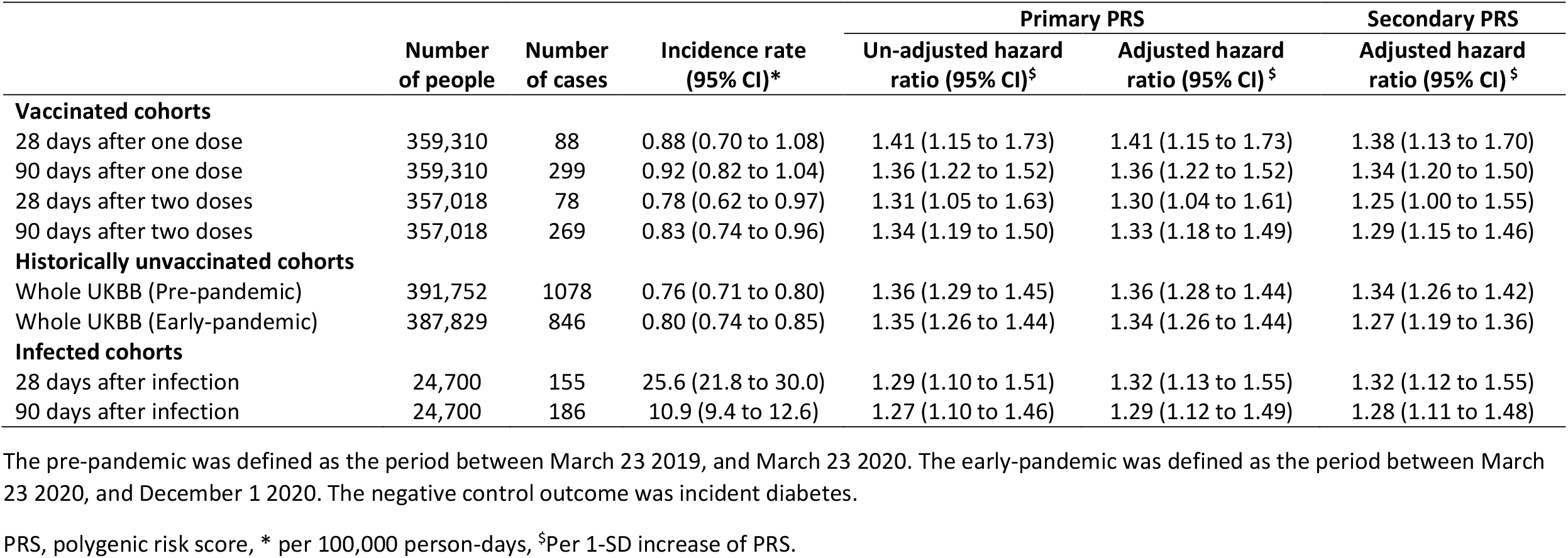
Association between the genetic score and incident venous thromboembolism in vaccinated and reference cohorts.

### Vaccination against covid-19

Vaccination status for UKBB participants was obtained from the linked primary care records provided by the two GP system suppliers: EMS and TPP (latest update: September 31, 2021). The clinical codes used for the first and second dose of the Covid-19 vaccines were “*1324681000000101”* and “*1324691000000104”* in EMS (SNOMED CT) and “*Y29e7*” and “*Y29e8*” in TPP (READ v3), respectively.

### Venous thromboembolism

VTE, including pulmonary embolism and deep vein thrombosis, was captured from linked hospital admission data from Hospital Episode Statistics, which contains all admissions in NHS hospitals in England. Mortality was ascertained from linked national death registry data. We used the earliest date of VTE diagnosis as the event date. ICD-10 codes used to identify VTE and death are given in the **sTable 3**.

**Table 3:** Exploratory analyses for different vaccine types.

| Cohorts | ChAdOx1 |  |  | BNT162b2 |  |  |
| --- | --- | --- | --- | --- | --- | --- |
|  | No. of cases/<br>people | Incidence rate<br>(95% CI)* | Adjusted hazard ratio<br>(95% CI) <sup>§</sup> | No. of cases/<br>people | Incidence rate<br>(95% CI)* | Adjusted hazard ratio<br>(95% CI) <sup>§</sup> |
| 28 days after one dose | 25 / 138059 | 0.65 (0.42 to 0.96) | 1.45 (0.99 to 2.13) | 33 / 83816 | 1.41 (0.97 to 1.98) | 1.38 (0.99 to 1.93) |
| 90 days after one dose | 94 / 138059 | 0.76 (0.61 to 0.93) | 1.63 (1.34 to 1.98) | 99 / 83816 | 1.31 (1.07 to 1.60) | 1.24 (1.02 to 1.50) |
| 28 days after two doses | 30 / 136002 | 0.79 (0.53 to 1.13) | 1.24 (0.88 to 1.77) | 26 / 80709 | 1.15 (0.75 to 1.69) | 1.20 (0.82 to 1.76) |
| 90 days after two doses | 100 / 136002 | 0.82 (0.67 to 1.00) | 1.53 (1.27 to 1.85) | 87 / 80709 | 1.20 (0.96 to 1.48) | 1.22 (1.00 to 1.51) |

\* per 100,000 person-days, <sup>§</sup>Per 1-SD increase of PRS.

### Statistical analyses

We used Cox proportional-hazards models to assess the associations between the PRS and VTE outcome. We computed hazard ratios (HR) and their 95% confidence intervals (CI) with adjustment for age (at the index date), sex, and genetic ancestry (quantified by the first ten principal components). To identify the high genetic risk group, we tested three cut-off quantiles of PRS separately, including upper tertile (top 33%), quintile (top 20%), and the top 5% with the lower 66% as the reference. To ensure sufficient statistical power, this analysis was only performed in the 90-days follow-up window. We evaluated the balance of baseline characteristics within each comparison pair according to a list of pre-specified covariates and adjusted for them in the Cox model if their absolute standardised mean difference was greater than 0.1. Considering varying VTE rates across the reference groups, we derived absolute risk increases (ARI) between high-risk and the reference PRS categories using the formula: (adjusted HR – 1) * cumulative incidence in the reference group.

We calculated HRs for diabetes as a negative control outcome to examine the specificity of the PRS and the likelihood of potential residual confounding. Diabetes was chosen with considerations that it is a well-developed disease phenotype and not biologically related to the VTE PRS. In a sub-cohort where the EMIS system provided the primary care data, and vaccine types were recorded, separate HRs were estimated among either ChAdOx1 or BNT162b2 vaccine recipients. Given that the heterologous prime-boost vaccination schedule in the UK is very uncommon^11^ (with <1% in our data), no specific analyses in this regard have been performed.

All the analyses were performed using PLINK1.9, QCTOOL v2, and R 4.1.2 software.

## Results

### Characteristics of vaccine recipients in UKBB

Of 380,822 UKBB participants eligible at the study entry (December 2, 2020), 378,662 (99.4%) and 376,416 (98.8%) received the first and second dose of Covid-19 vaccines, respectively, until the study end date (September 31, 2021) (**Figure 1**). For the one-dose cohort, the mean age was 69.05 years (standard deviation 8.04), and 160,327 (44.6%) were male (**Table 1**). A similar demographic profile was observed for the two-dose cohort (**Table 1**). The PRS approximated a normal distribution within each cohort (**sFigure 1**).

**Figure 1:**
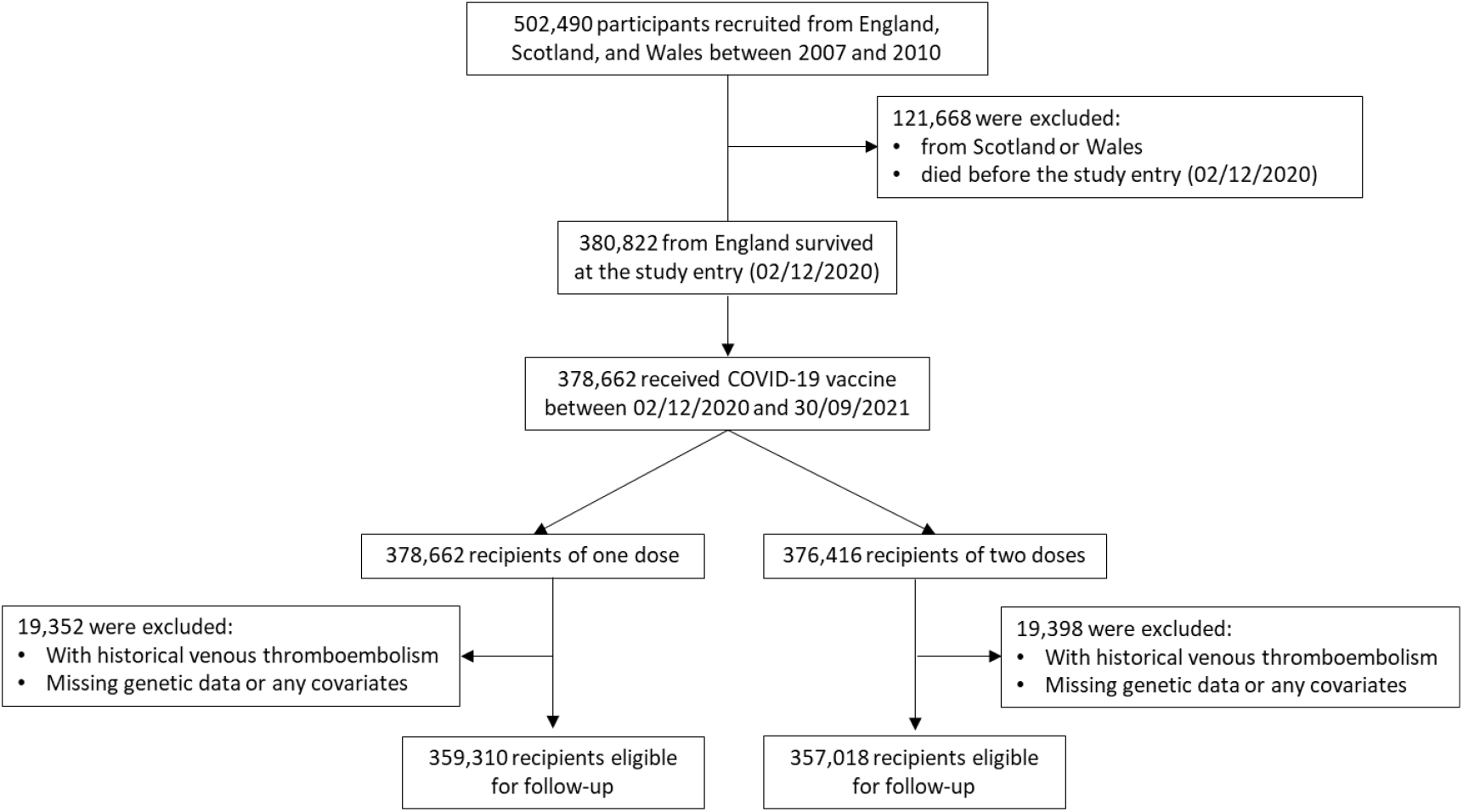
Flow chart of the study selection process.

### Association of the PRS with incident VTE

During the follow-up periods, 88 and 299 individuals developed VTE within 28 and 90 days after first-dose vaccination (**Table 2**), equivalent to an incidence rate of 0.88 (95% CI 0.70 to 1.08) and 0.92 (95% CI 0.82 to 1.04) per 100,000 person-days. The unadjusted and adjusted HRs for VTE associated with the primary PRS were similar, with the latter being 1.41 (95% 1.15 to 1.73) per 1-SD increase in PRS (1-SD-PRS) over 28-day follow-up and 1.36 (95% 1.22 to 1.52) over 90-days. The association between the PRS value and risk of VTE appears to be monotonic in nature (**sFigure 2**). After the second dose vaccination, the association between PRS and VTE was slightly attenuated (HR: 1.30 (95% 1.04 to 1.61) per 1-SD-PRS and 1.33 (95% 1.18 to 1.49) in the 28- and 90-days’ follow-up window, respectively) (**Table 2**). Although there was a seemingly inverted U-shaped relationship between the PRS and estimate of VTE risk following the second dose of vaccine, wide confidence intervals limit the reliability of this finding.

The observed rates and effect sizes of the observed associations were similar when comparing the vaccinated and historical (unvaccinated) cohorts. Also, although absolute incidence rates of VTE in the infected cohort were substantially higher than those in other cohorts, the PRS-VTE association persisted. A sensitivity analysis using an alternative PRS found similar although slightly weaker associations (**Table 2**).

Finally, no associations were observed for our proposed negative control outcome: the HR between PRS and incident diabetes was 1.02 (95% 0.98 to 1.06) in the pre-pandemic and 0.98 (95% 0.93 to 1.04) in the early pandemic period (**sTable 4**).

### Identification of high-risk group

Figure 2 presents HRs and ARI for VTE across three pre-defined high-risk categories. Briefly, relative risks increased with cut-offs from 33% to 5%, corresponding to HRs ranging from 1.67 (95% 1.33 to 2.09) to 2.10 (95% 1.39 to 3.18) in the one- and from 1.66 (95% 1.30 to 2.11) to 1.97 (95% 1.26 to 3.09) in the two-dose cohorts. Also, there was a linear increasing trend for absolute risk differences, with ARI of 0.45 (95% 0.22 to 0.74) to 0.76 (95% .27 to 1.51) and 0.40 (95% 0.19 to 0.67) to 0.59 (95% 0.16 to 1.28) in the one- and two-dose cohort respectively.

**Figure 2:**
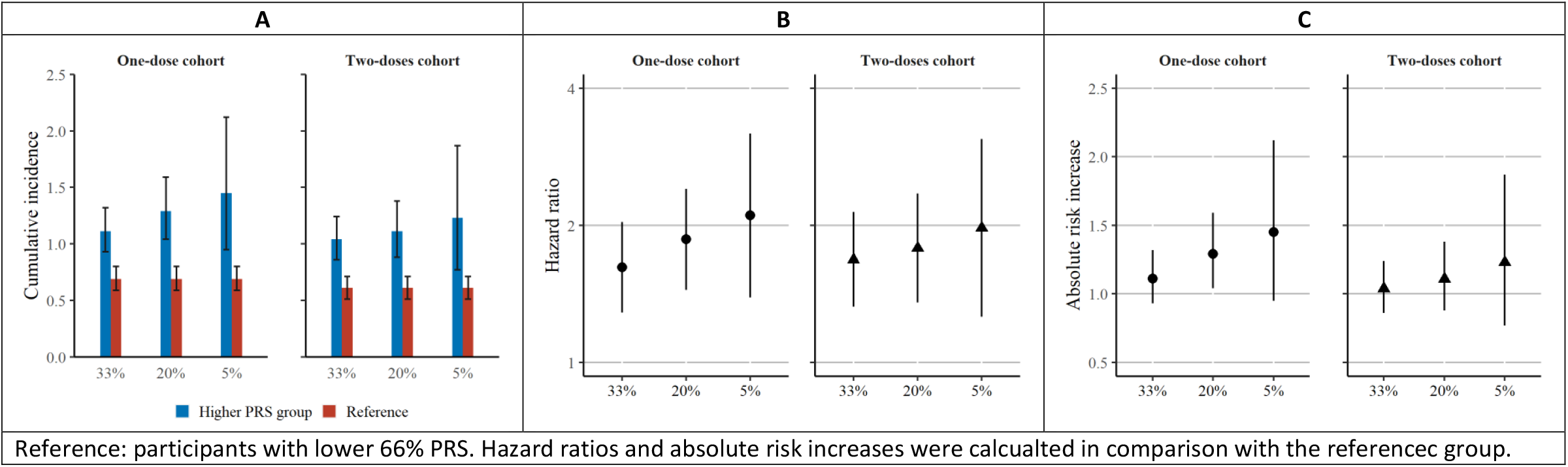
90-day cumulative incidence (A), hazard ratios (B) and absolute risk increases (C) of three pre-defined high genetic risk groups vs the reference.

### Different vaccine types

Among 221,875 recipients with vaccine-type information available (138,059 received ChAdOx1 and 83,816 received BNT162b2), the observed PRS-VTE associations were similar across each dose and follow-up window: HR ranged from 1.24 (95% CI 0.88 to 1.77) to 1.63 (95% CI 1.34 to 1.98) in ChAdOx1 vaccinated cohorts, and from 1.20 (95% CI 0.82 to 1.76) to 1.38 (95% CI 0.99 to 1.93) in BNT162b2 vaccinated people (**Table 3**). Noticeably, the background VTE incidence rates in BNT162b2 vaccinated cohorts were almost doubly higher than those in the ChAdOx1 vaccinated one, which was expected given that the former vaccine was approved earlier in the UK and prioritised for older and more vulnerable populations.^12^

## Discussion

Our study showed that a PRS for VTE could identify people at increased risk of VTE within 28 or 90 days after receiving one or two doses of Covid-19 vaccines. Furthermore, the strength of the PRS association to post-Covid-19-vaccination VTE was similar to that seen for VTE before Covid-19 vaccination rollout.

The PRS used in the present study was developed and validated by Klarin .et al. Such study found a 2.5 to 3-fold increased risk of VTE associated with the highest 5% of the score in both case-control and prospective cohort study settings.^9^ Recently, Marston et al. tested the performance of the PRS among cardiometabolic disease patients to predict VTE and observed a similar magnitude of effect (2.7-fold for top 33 % vs bottom 66%).^13^ Despite being aligned with these findings, the PRS-VTE associations estimated in our study were consistently weaker than the previously reported ones even after the incorporation of the two clinically validated variants, possibly due to the discrepancies in defining VTE phenotypes between the original score deviation and this validation study. Also, because our cohort only consisted of VTE naïve and relatively older participants, those with higher genetic risk might have had a VTE in their earlier age and thus been excluded. As expected, our PRS was not associated with the proposed negative control outcome (incident diabetes), to some extent, demonstrating its specificity for VTE prediction.

The results of this study support several noteworthy conclusions. First, our data showed that individuals’ genetic susceptibility to VTE was a risk factor for VTE among the Covid-19 vaccinated population. Second, this genetic risk was independent of traditional risk factors such as old age, obesity and comorbidity, as indicated by no associations between the PRS and baseline characteristics (**Table 1**). Third, by designing a historical comparison arm in the same population, our data suggest that clinically significant interactions between individuals’ genetic background and Covid-19 vaccination are unlikely, which has particular implications for patients with hereditary VTE predisposing traits who are hesitant to be vaccinated because of concerns regarding related recent vaccine safety signals. Fourth, we identified 5% of people with more than 2-fold higher VTE risk by using this genetic score, it should be of public health relevance and can inform potential intervention policies given the absolute size of Covid-19 vaccinated population. Our analyses have some potential limitations. First, VTE often presents variable clinical manifestations with challenging differential diagnoses such as myocardial infarction and congestive heart failure.^2^ Consequently, identification of VTE in a real-world setting is likely subject to information bias, which typically drives risk estimates towards the null. Second, we were not able to generate a parallel unvaccinated comparison group because more than 99% of UKBB participants had been vaccinated. However, we constructed a historical comparison cohort with similar characteristics to those vaccinated. Also, given the relatively short follow-up after vaccination, the long-term impact of the genetic factor remains to be determined. Third, although we also constructed a secondary PRS for VTE, the weights of each included SNP have not been previously validated, and their utility in a PRS remains unknown.

Opportunely, it conferred consistent results as the primary PRS did, likely due to the fact that both PRSs included the factor V Leiden p.R506Q and prothrombin G20210A variants which are known causes of inherited thrombophilia predisposing to acute thrombotic syndromes^14,15^. Fourth, the HR estimates for each vaccine type should be considered exploratory in nature due to evident differences in the baseline risk for VTE seen between people vaccinated with the two vaccines. Last, the generalizability of our findings should be tested in more diverse ethnic populations as more integrated data sources containing in-depth genetic, vaccination, and health information becomes available.

This study benefits from the use of a large prospective cohort with comprehensive genetic, Covid-19 vaccination, Covid-19 infection status and VTE phenotype data linked at the individual level, the application of the state-of-the-art PRS, and robust analytic methods by designing multiple comparison groups and a negative control outcome. To our knowledge, this is the first study to show that individuals’ who developed post-Covid19-vaccination VTE had a genetic predisposition to VTE, and that the association between the genetic risk factors and post-Covid19-vaccination VTE is similar to the association with conventional VTE.

## Conclusions

A published PRS for VTE, constructed using common genetic variants with small effects on VTE, was associated with increased VTE risk following Covid-19 vaccination. This association was similar to that seen historically, both in pre-pandemic times and during the first year of the Covid-19 pandemic, before vaccines were available. Our data do not support a clinically meaningful interplay between genetic predisposition and Covid-19 vaccines on the occurrence of VTE events. These findings suggest that the clinical management of VTE among the vaccinated population should not be disturbed by the concern of gene-vaccine interaction, and that people at high genetic risk of VTE such as those with inherited thrombophilia might have a modest excess risk of VTE occurrence following vaccination.

## Disclaimer

The views expressed in this article are the personal views of the author(s) and may not be understood or quoted as being made on behalf of or reflecting the position of the regulatory agency/agencies or organisations with which the author(s) is/are employed/affiliated.

## Data Availability

Bonafide researchers can apply to use the UK Biobank dataset by registering and applying at http://ukbiobank.ac.uk/register-apply/. The datasets generated and/or analysed during the current study are available from the corresponding author on reasonable request.

## Ethical approval

All participants provided written informed consent at the UKBB cohort recruitment. This study received ethical approval from UKBB Ethics Advisory Committee (EAC) and was performed under the application of 65397.

## Transparency declaration

The lead author affirms that this manuscript is an honest, accurate, and transparent account of the study being reported; that no important aspects of the study have been omitted.

## Contributors

D.P.A., J.Q.X., D.G were responsible for the study design. J.Q.X. did the data analyses, and A.P.U. checked the statistical codes. J.Q.X. and D.P.A. drafted the manuscript, and all co-authors reviewed and approved it for submission.

## Supplementary materials

**sFigure 1:**
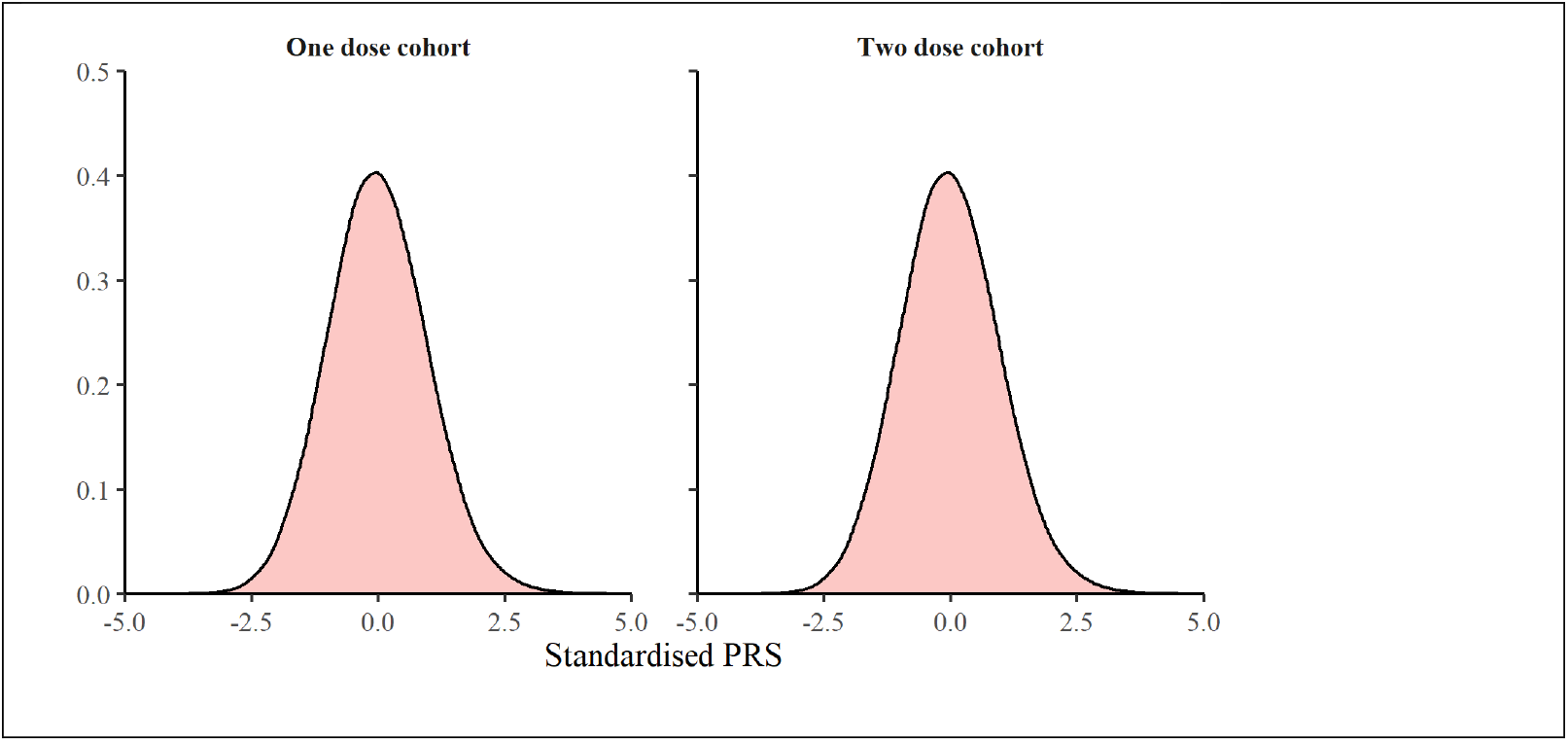
Distribution of polygenic risk score for venous thromboembolism.

**sFigure 2:**
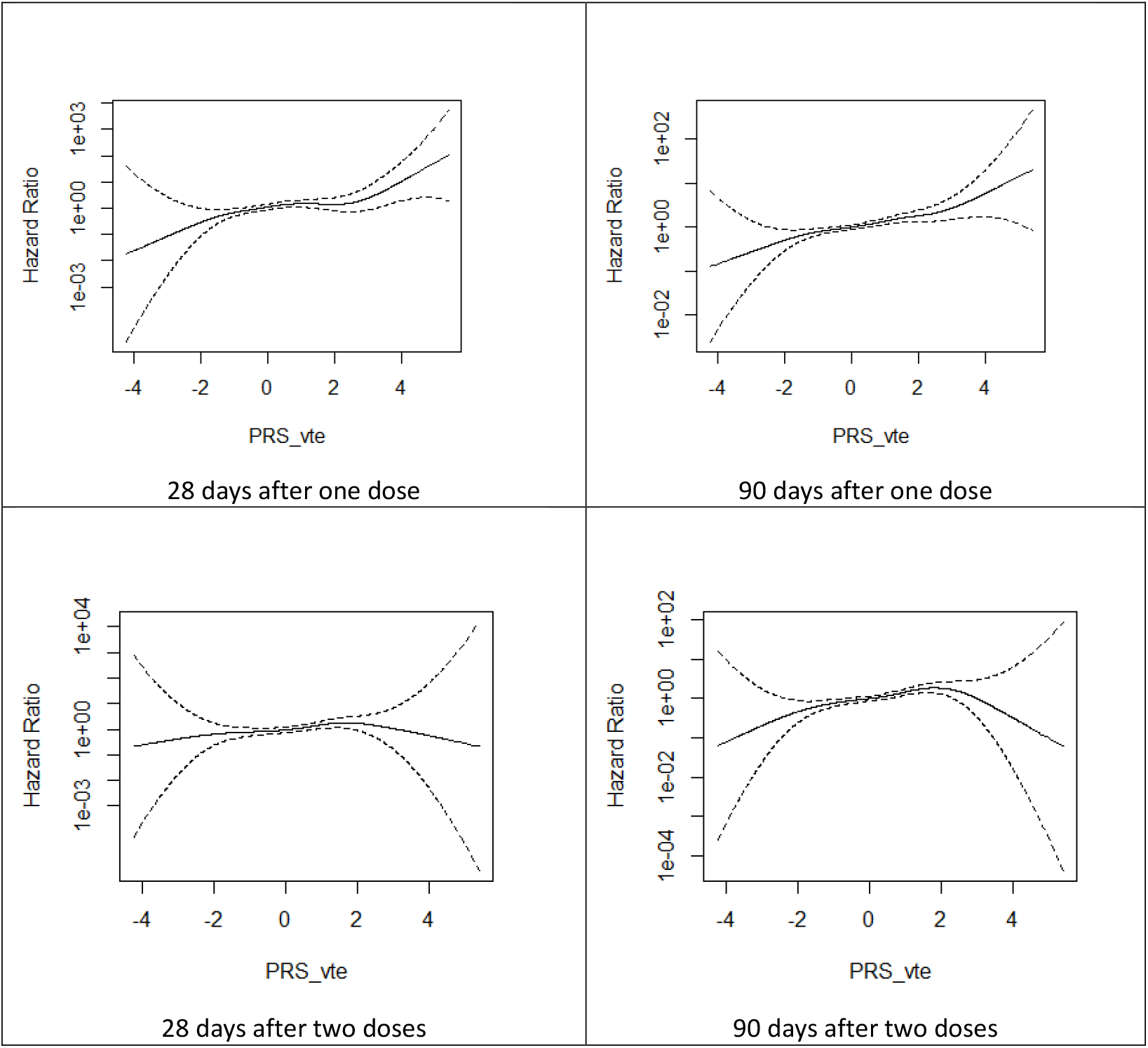
The relationship between the PRS value and the risk (hazard ratio) of VTE.

**sTable 1:**
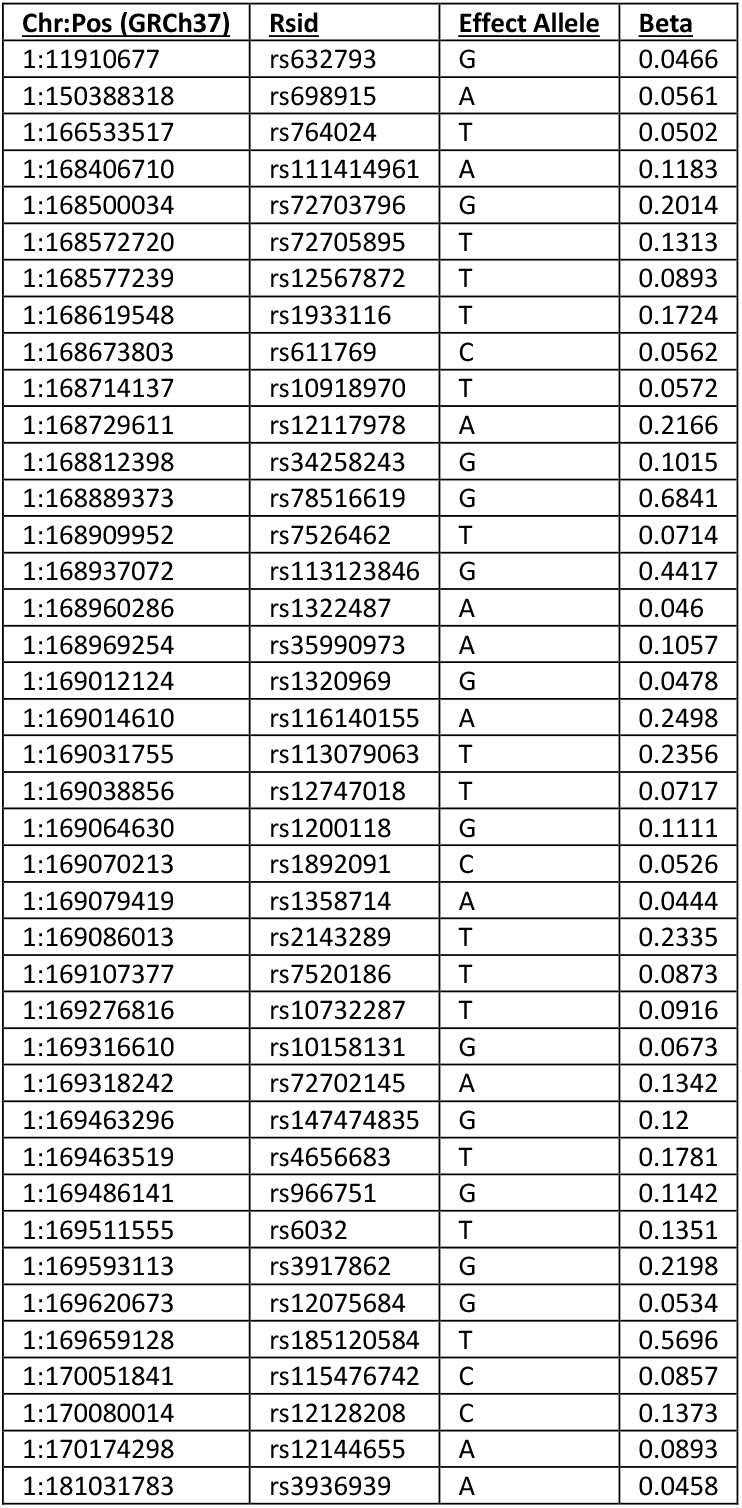
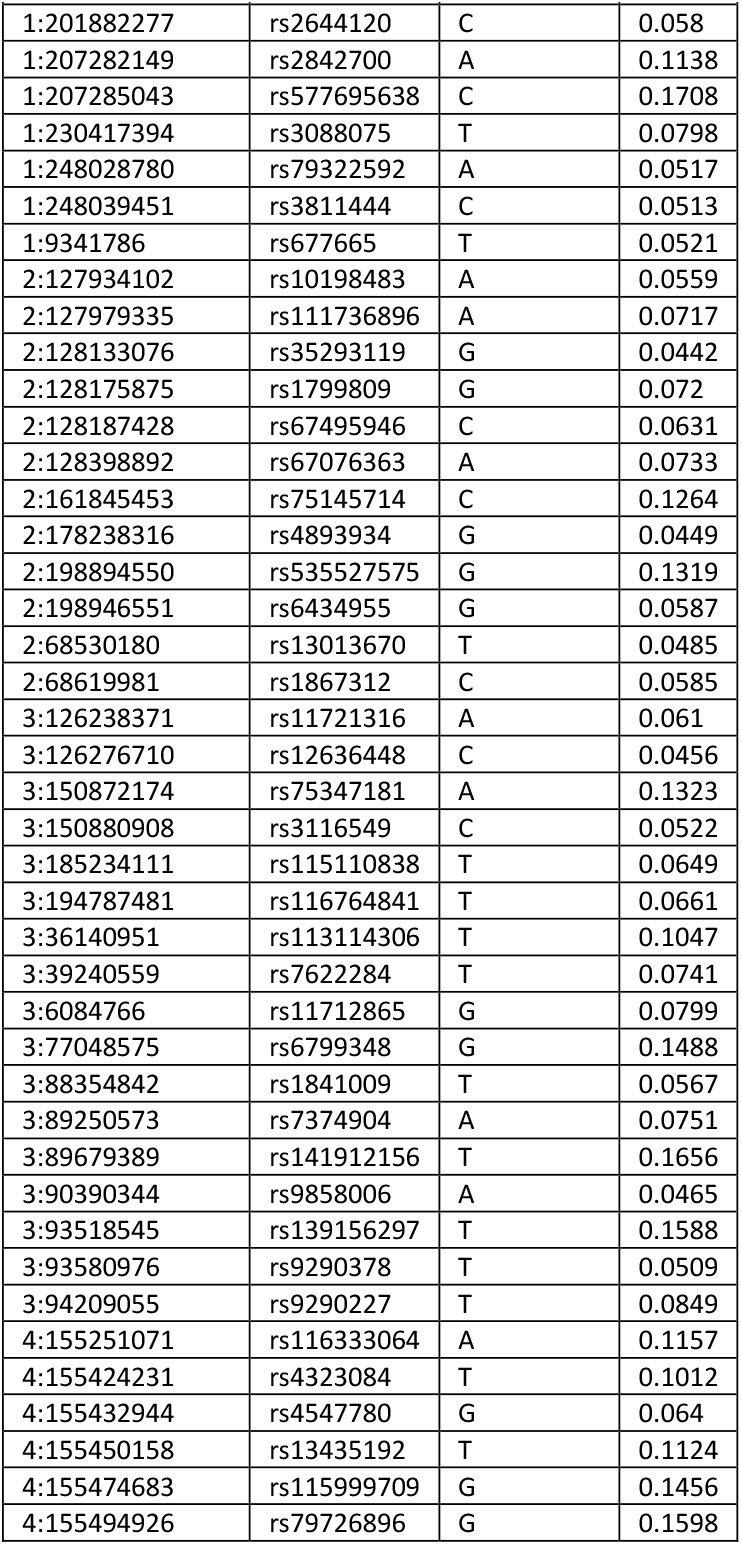
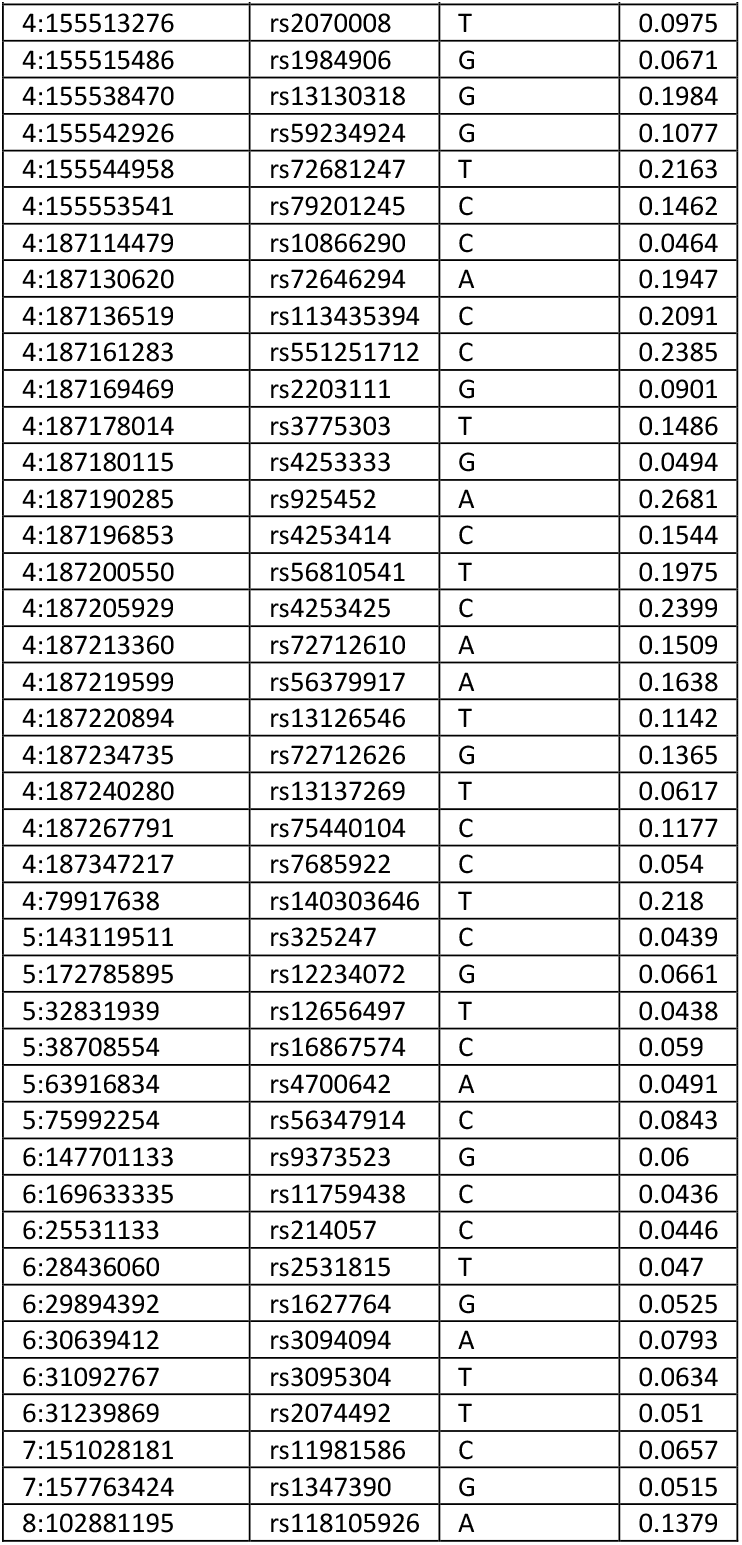
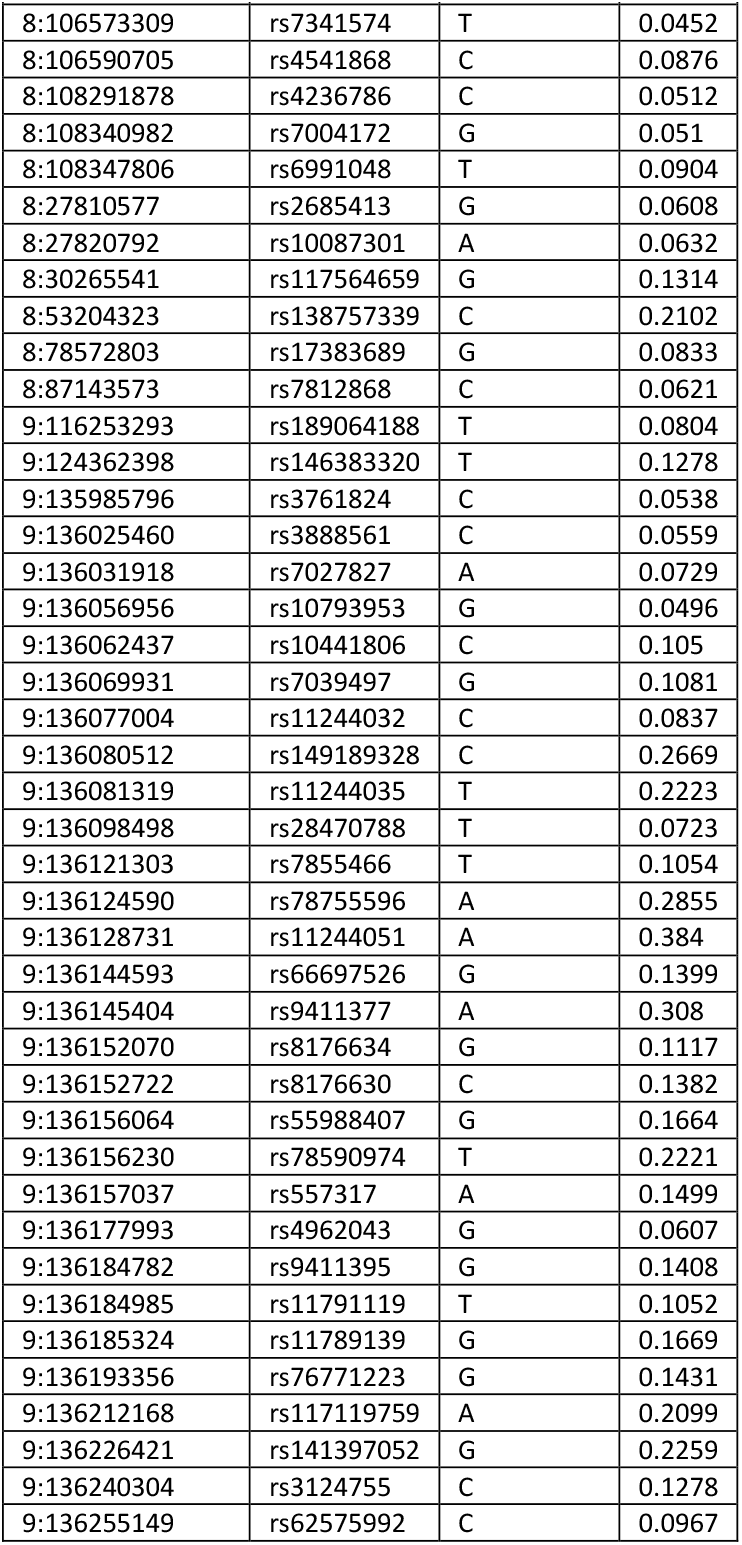
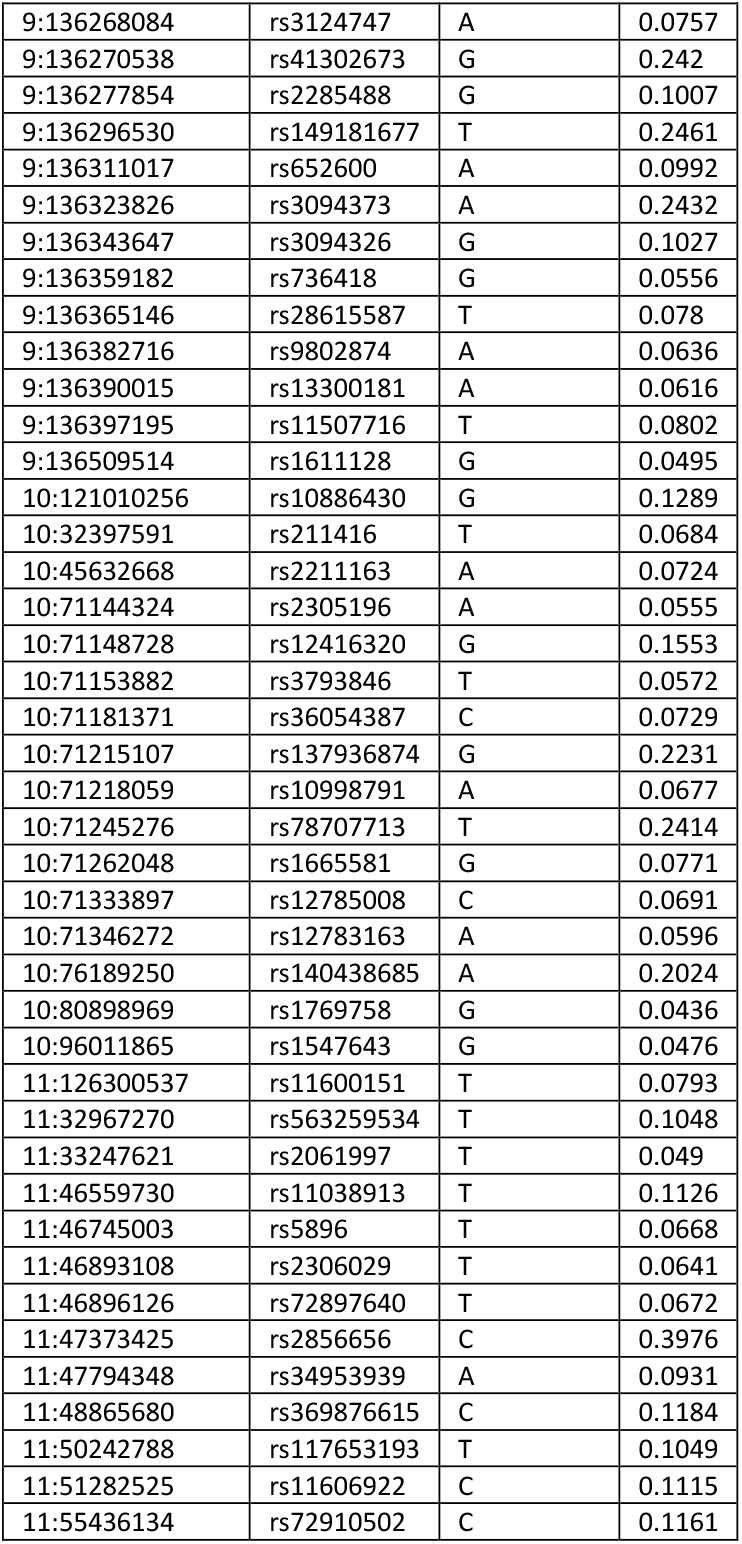
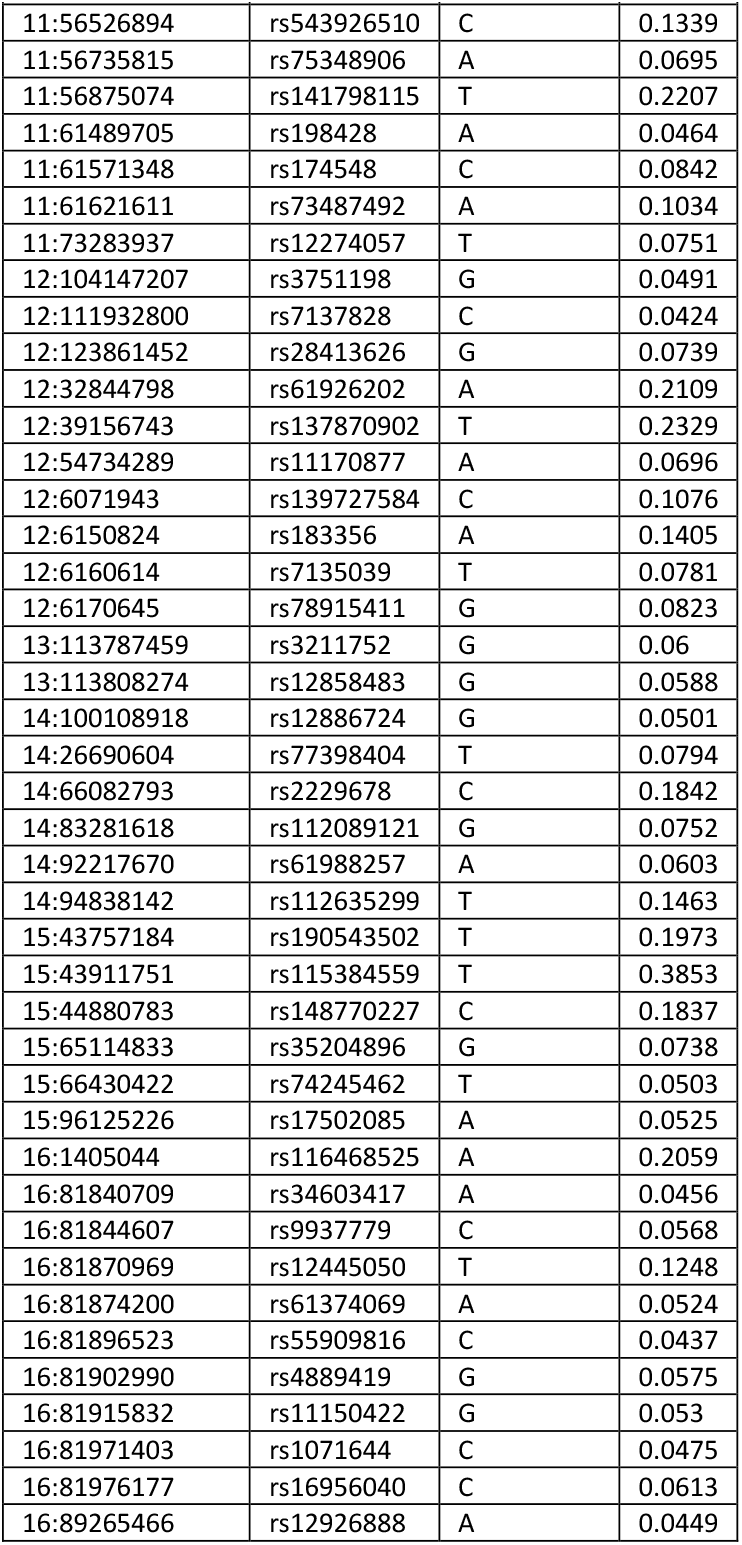
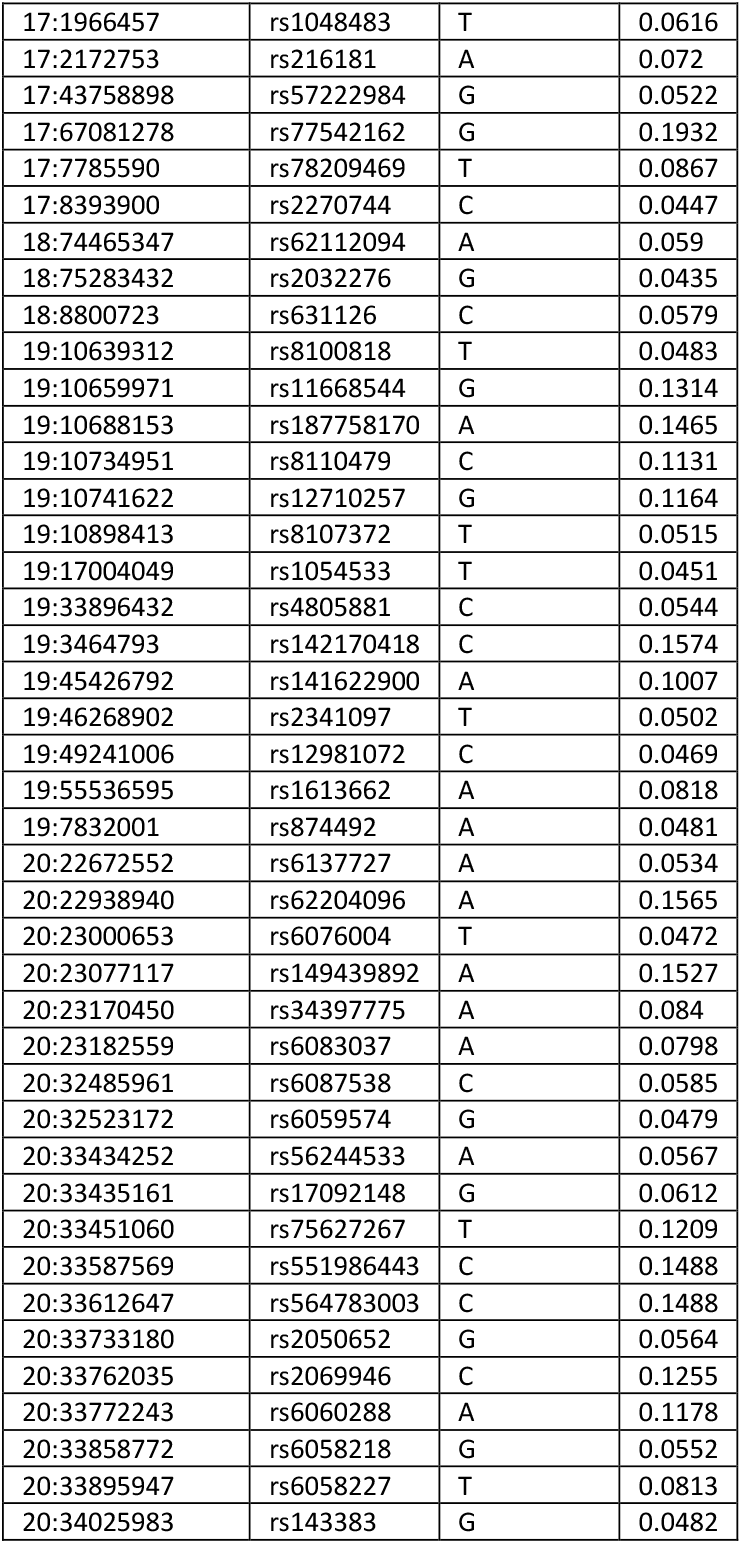
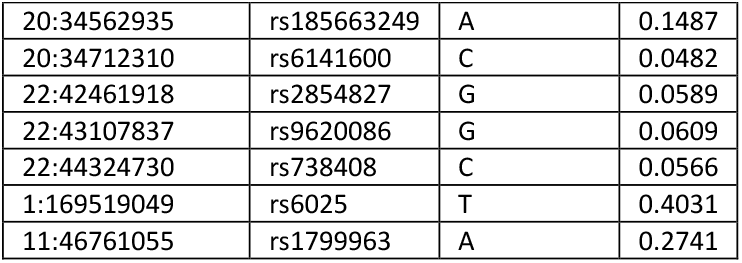
Summary statistics for candidate SNPs used for constructing the main polygenic risk score (Main PRS_VTE).

| **Chr:Pos (GRCh37)** | **Rsid** | **Effect Allele** | **Beta** |
| --- | --- | --- | --- |
| 1:11910677 | rs632793 | G | 0.0466 |
| 1:150388318 | rs698915 | A | 0.0561 |
| 1:166533517 | rs764024 | T | 0.0502 |
| 1:168406710 | rs111414961 | A | 0.1183 |
| 1:168500034 | rs72703796 | G | 0.2014 |
| 1:168572720 | rs72705895 | T | 0.1313 |
| 1:168577239 | rs12567872 | T | 0.0893 |
| 1:168619548 | rs1933116 | T | 0.1724 |
| 1:168673803 | rs611769 | C | 0.0562 |
| 1:168714137 | rs10918970 | T | 0.0572 |
| 1:168729611 | rs12117978 | A | 0.2166 |
| 1:168812398 | rs34258243 | G | 0.1015 |
| 1:168889373 | rs78516619 | G | 0.6841 |
| 1:168909952 | rs7526462 | T | 0.0714 |
| 1:168937072 | rs113123846 | G | 0.4417 |
| 1:168960286 | rs1322487 | A | 0.046 |
| 1:168969254 | rs35990973 | A | 0.1057 |
| 1:169012124 | rs1320969 | G | 0.0478 |
| 1:169014610 | rs116140155 | A | 0.2498 |
| 1:169031755 | rs113079063 | T | 0.2356 |
| 1:169038856 | rs12747018 | T | 0.0717 |
| 1:169064630 | rs1200118 | G | 0.1111 |
| 1:169070213 | rs1892091 | C | 0.0526 |
| 1:169079419 | rs1358714 | A | 0.0444 |
| 1:169086013 | rs2143289 | T | 0.2335 |
| 1:169107377 | rs7520186 | T | 0.0873 |
| 1:169276816 | rs10732287 | T | 0.0916 |
| 1:169316610 | rs10158131 | G | 0.0673 |
| 1:169318242 | rs72702145 | A | 0.1342 |
| 1:169463296 | rs147474835 | G | 0.12 |
| 1:169463519 | rs4656683 | T | 0.1781 |
| 1:169486141 | rs966751 | G | 0.1142 |
| 1:169511555 | rs6032 | T | 0.1351 |
| 1:169593113 | rs3917862 | G | 0.2198 |
| 1:169620673 | rs12075684 | G | 0.0534 |
| 1:169659128 | rs185120584 | T | 0.5696 |
| 1:170051841 | rs115476742 | C | 0.0857 |
| 1:170080014 | rs12128208 | C | 0.1373 |
| 1:170174298 | rs12144655 | A | 0.0893 |
| 1:181031783 | rs3936939 | A | 0.0458 |

| Chr:Pos (GRCh37) | Rsid | Effect Allele | Beta |
| --- | --- | --- | --- |
| 1:201882277 | rs2644120 | C | 0.058 |
| 1:207282149 | rs2842700 | A | 0.1138 |
| 1:207285043 | rs577695638 | C | 0.1708 |
| 1:230417394 | rs3088075 | T | 0.0798 |
| 1:248028780 | rs79322592 | A | 0.0517 |
| 1:248039451 | rs3811444 | C | 0.0513 |
| 1:9341786 | rs677665 | T | 0.0521 |
| 2:127934102 | rs10198483 | A | 0.0559 |
| 2:127979335 | rs111736896 | A | 0.0717 |
| 2:128133076 | rs35293119 | G | 0.0442 |
| 2:128175875 | rs1799809 | G | 0.072 |
| 2:128187428 | rs67495946 | C | 0.0631 |
| 2:128398892 | rs67076363 | A | 0.0733 |
| 2:161845453 | rs75145714 | C | 0.1264 |
| 2:178238316 | rs4893934 | G | 0.0449 |
| 2:198894550 | rs535527575 | G | 0.1319 |
| 2:198946551 | rs6434955 | G | 0.0587 |
| 2:68530180 | rs13013670 | T | 0.0485 |
| 2:68619981 | rs1867312 | C | 0.0585 |
| 3:126238371 | rs11721316 | A | 0.061 |
| 3:126276710 | rs12636448 | C | 0.0456 |
| 3:150872174 | rs75347181 | A | 0.1323 |
| 3:150880908 | rs3116549 | C | 0.0522 |
| 3:185234111 | rs115110838 | T | 0.0649 |
| 3:194787481 | rs116764841 | T | 0.0661 |
| 3:36140951 | rs113114306 | T | 0.1047 |
| 3:39240559 | rs7622284 | T | 0.0741 |
| 3:6084766 | rs11712865 | G | 0.0799 |
| 3:77048575 | rs6799348 | G | 0.1488 |
| 3:88354842 | rs1841009 | T | 0.0567 |
| 3:89250573 | rs7374904 | A | 0.0751 |
| 3:89679389 | rs141912156 | T | 0.1656 |
| 3:90390344 | rs9858006 | A | 0.0465 |
| 3:93518545 | rs139156297 | T | 0.1588 |
| 3:93580976 | rs9290378 | T | 0.0509 |
| 3:94209055 | rs9290227 | T | 0.0849 |
| 4:155251071 | rs116333064 | A | 0.1157 |
| 4:155424231 | rs4323084 | T | 0.1012 |
| 4:155432944 | rs4547780 | G | 0.064 |
| 4:155450158 | rs13435192 | T | 0.1124 |
| 4:155474683 | rs115999709 | G | 0.1456 |
| 4:155494926 | rs79726896 | G | 0.1598 |
| 4:155513276 | rs2070008 | T | 0.0975 |
| 4:155515486 | rs1984906 | G | 0.0671 |
| 4:155538470 | rs13130318 | G | 0.1984 |
| 4:155542926 | rs59234924 | G | 0.1077 |
| 4:155544958 | rs72681247 | T | 0.2163 |
| 4:155553541 | rs79201245 | C | 0.1462 |
| 4:187114479 | rs10866290 | C | 0.0464 |
| 4:187130620 | rs72646294 | A | 0.1947 |
| 4:187136519 | rs113435394 | C | 0.2091 |
| 4:187161283 | rs551251712 | C | 0.2385 |
| 4:187169469 | rs2203111 | G | 0.0901 |
| 4:187178014 | rs3775303 | T | 0.1486 |
| 4:187180115 | rs4253333 | G | 0.0494 |
| 4:187190285 | rs925452 | A | 0.2681 |
| 4:187196853 | rs4253414 | C | 0.1544 |
| 4:187200550 | rs56810541 | T | 0.1975 |
| 4:187205929 | rs4253425 | C | 0.2399 |
| 4:187213360 | rs72712610 | A | 0.1509 |
| 4:187219599 | rs56379917 | A | 0.1638 |
| 4:187220894 | rs13126546 | T | 0.1142 |
| 4:187234735 | rs72712626 | G | 0.1365 |
| 4:187240280 | rs13137269 | T | 0.0617 |
| 4:187267791 | rs75440104 | C | 0.1177 |
| 4:187347217 | rs7685922 | C | 0.054 |
| 4:79917638 | rs140303646 | T | 0.218 |
| 5:143119511 | rs325247 | C | 0.0439 |
| 5:172785895 | rs12234072 | G | 0.0661 |
| 5:32831939 | rs12656497 | T | 0.0438 |
| 5:38708554 | rs16867574 | C | 0.059 |
| 5:63916834 | rs4700642 | A | 0.0491 |
| 5:75992254 | rs56347914 | C | 0.0843 |
| 6:147701133 | rs9373523 | G | 0.06 |
| 6:169633335 | rs11759438 | C | 0.0436 |
| 6:25531133 | rs214057 | C | 0.0446 |
| 6:28436060 | rs2531815 | T | 0.047 |
| 6:29894392 | rs1627764 | G | 0.0525 |
| 6:30639412 | rs3094094 | A | 0.0793 |
| 6:31092767 | rs3095304 | T | 0.0634 |
| 6:31239869 | rs2074492 | T | 0.051 |
| 7:151028181 | rs11981586 | C | 0.0657 |
| 7:157763424 | rs1347390 | G | 0.0515 |
| 8:102881195 | rs118105926 | A | 0.1379 |
| 8:106573309 | rs7341574 | T | 0.0452 |
| 8:106590705 | rs4541868 | C | 0.0876 |
| 8:108291878 | rs4236786 | C | 0.0512 |
| 8:108340982 | rs7004172 | G | 0.051 |
| 8:108347806 | rs6991048 | T | 0.0904 |
| 8:27810577 | rs2685413 | G | 0.0608 |
| 8:27820792 | rs10087301 | A | 0.0632 |
| 8:30265541 | rs117564659 | G | 0.1314 |
| 8:53204323 | rs138757339 | C | 0.2102 |
| 8:78572803 | rs17383689 | G | 0.0833 |
| 8:87143573 | rs7812868 | C | 0.0621 |
| 9:116253293 | rs189064188 | T | 0.0804 |
| 9:124362398 | rs146383320 | T | 0.1278 |
| 9:135985796 | rs3761824 | C | 0.0538 |
| 9:136025460 | rs3888561 | C | 0.0559 |
| 9:136031918 | rs7027827 | A | 0.0729 |
| 9:136056956 | rs10793953 | G | 0.0496 |
| 9:136062437 | rs10441806 | C | 0.105 |
| 9:136069931 | rs7039497 | G | 0.1081 |
| 9:136077004 | rs11244032 | C | 0.0837 |
| 9:136080512 | rs149189328 | C | 0.2669 |
| 9:136081319 | rs11244035 | T | 0.2223 |
| 9:136098498 | rs28470788 | T | 0.0723 |
| 9:136121303 | rs7855466 | T | 0.1054 |
| 9:136124590 | rs78755596 | A | 0.2855 |
| 9:136128731 | rs11244051 | A | 0.384 |
| 9:136144593 | rs66697526 | G | 0.1399 |
| 9:136145404 | rs9411377 | A | 0.308 |
| 9:136152070 | rs8176634 | G | 0.1117 |
| 9:136152722 | rs8176630 | C | 0.1382 |
| 9:136156064 | rs55988407 | G | 0.1664 |
| 9:136156230 | rs78590974 | T | 0.2221 |
| 9:136157037 | rs557317 | A | 0.1499 |
| 9:136177993 | rs4962043 | G | 0.0607 |
| 9:136184782 | rs9411395 | G | 0.1408 |
| 9:136184985 | rs11791119 | T | 0.1052 |
| 9:136185324 | rs11789139 | G | 0.1669 |
| 9:136193356 | rs76771223 | G | 0.1431 |
| 9:136212168 | rs117119759 | A | 0.2099 |
| 9:136226421 | rs141397052 | G | 0.2259 |
| 9:136240304 | rs3124755 | C | 0.1278 |
| 9:136255149 | rs62575992 | C | 0.0967 |
| 9:136268084 | rs3124747 | A | 0.0757 |
| 9:136270538 | rs41302673 | G | 0.242 |
| 9:136277854 | rs2285488 | G | 0.1007 |
| 9:136296530 | rs149181677 | T | 0.2461 |
| 9:136311017 | rs652600 | A | 0.0992 |
| 9:136323826 | rs3094373 | A | 0.2432 |
| 9:136343647 | rs3094326 | G | 0.1027 |
| 9:136359182 | rs736418 | G | 0.0556 |
| 9:136365146 | rs28615587 | T | 0.078 |
| 9:136382716 | rs9802874 | A | 0.0636 |
| 9:136390015 | rs13300181 | A | 0.0616 |
| 9:136397195 | rs11507716 | T | 0.0802 |
| 9:136509514 | rs1611128 | G | 0.0495 |
| 10:121010256 | rs10886430 | G | 0.1289 |
| 10:32397591 | rs211416 | T | 0.0684 |
| 10:45632668 | rs2211163 | A | 0.0724 |
| 10:71144324 | rs2305196 | A | 0.0555 |
| 10:71148728 | rs12416320 | G | 0.1553 |
| 10:71153882 | rs3793846 | T | 0.0572 |
| 10:71181371 | rs36054387 | C | 0.0729 |
| 10:71215107 | rs137936874 | G | 0.2231 |
| 10:71218059 | rs10998791 | A | 0.0677 |
| 10:71245276 | rs78707713 | T | 0.2414 |
| 10:71262048 | rs1665581 | G | 0.0771 |
| 10:71333897 | rs12785008 | C | 0.0691 |
| 10:71346272 | rs12783163 | A | 0.0596 |
| 10:76189250 | rs140438685 | A | 0.2024 |
| 10:80898969 | rs1769758 | G | 0.0436 |
| 10:96011865 | rs1547643 | G | 0.0476 |
| 11:126300537 | rs11600151 | T | 0.0793 |
| 11:32967270 | rs563259534 | T | 0.1048 |
| 11:33247621 | rs2061997 | T | 0.049 |
| 11:46559730 | rs11038913 | T | 0.1126 |
| 11:46745003 | rs5896 | T | 0.0668 |
| 11:46893108 | rs2306029 | T | 0.0641 |
| 11:46896126 | rs72897640 | T | 0.0672 |
| 11:47373425 | rs2856656 | C | 0.3976 |
| 11:47794348 | rs34953939 | A | 0.0931 |
| 11:48865680 | rs369876615 | C | 0.1184 |
| 11:50242788 | rs117653193 | T | 0.1049 |
| 11:51282525 | rs11606922 | C | 0.1115 |
| 11:55436134 | rs72910502 | C | 0.1161 |
| 11:56526894 | rs543926510 | C | 0.1339 |
| 11:56735815 | rs75348906 | A | 0.0695 |
| 11:56875074 | rs141798115 | T | 0.2207 |
| 11:61489705 | rs198428 | A | 0.0464 |
| 11:61571348 | rs174548 | C | 0.0842 |
| 11:61621611 | rs73487492 | A | 0.1034 |
| 11:73283937 | rs12274057 | T | 0.0751 |
| 12:104147207 | rs3751198 | G | 0.0491 |
| 12:111932800 | rs7137828 | C | 0.0424 |
| 12:123861452 | rs28413626 | G | 0.0739 |
| 12:32844798 | rs61926202 | A | 0.2109 |
| 12:39156743 | rs137870902 | T | 0.2329 |
| 12:54734289 | rs11170877 | A | 0.0696 |
| 12:6071943 | rs139727584 | C | 0.1076 |
| 12:6150824 | rs183356 | A | 0.1405 |
| 12:6160614 | rs7135039 | T | 0.0781 |
| 12:6170645 | rs78915411 | G | 0.0823 |
| 13:113787459 | rs3211752 | G | 0.06 |
| 13:113808274 | rs12858483 | G | 0.0588 |
| 14:100108918 | rs12886724 | G | 0.0501 |
| 14:26690604 | rs77398404 | T | 0.0794 |
| 14:66082793 | rs2229678 | C | 0.1842 |
| 14:83281618 | rs112089121 | G | 0.0752 |
| 14:92217670 | rs61988257 | A | 0.0603 |
| 14:94838142 | rs112635299 | T | 0.1463 |
| 15:43757184 | rs190543502 | T | 0.1973 |
| 15:43911751 | rs115384559 | T | 0.3853 |
| 15:44880783 | rs148770227 | C | 0.1837 |
| 15:65114833 | rs35204896 | G | 0.0738 |
| 15:66430422 | rs74245462 | T | 0.0503 |
| 15:96125226 | rs17502085 | A | 0.0525 |
| 16:1405044 | rs116468525 | A | 0.2059 |
| 16:81840709 | rs34603417 | A | 0.0456 |
| 16:81844607 | rs9937779 | C | 0.0568 |
| 16:81870969 | rs12445050 | T | 0.1248 |
| 16:81874200 | rs61374069 | A | 0.0524 |
| 16:81896523 | rs55909816 | C | 0.0437 |
| 16:81902990 | rs4889419 | G | 0.0575 |
| 16:81915832 | rs11150422 | G | 0.053 |
| 16:81971403 | rs1071644 | C | 0.0475 |
| 16:81976177 | rs16956040 | C | 0.0613 |
| 16:89265466 | rs12926888 | A | 0.0449 |
| 17:1966457 | rs1048483 | T | 0.0616 |
| 17:2172753 | rs216181 | A | 0.072 |
| 17:43758898 | rs57222984 | G | 0.0522 |
| 17:67081278 | rs77542162 | G | 0.1932 |
| 17:7785590 | rs78209469 | T | 0.0867 |
| 17:8393900 | rs2270744 | C | 0.0447 |
| 18:74465347 | rs62112094 | A | 0.059 |
| 18:75283432 | rs2032276 | G | 0.0435 |
| 18:8800723 | rs631126 | C | 0.0579 |
| 19:10639312 | rs8100818 | T | 0.0483 |
| 19:10659971 | rs11668544 | G | 0.1314 |
| 19:10688153 | rs187758170 | A | 0.1465 |
| 19:10734951 | rs8110479 | C | 0.1131 |
| 19:10741622 | rs12710257 | G | 0.1164 |
| 19:10898413 | rs8107372 | T | 0.0515 |
| 19:17004049 | rs1054533 | T | 0.0451 |
| 19:33896432 | rs4805881 | C | 0.0544 |
| 19:3464793 | rs142170418 | C | 0.1574 |
| 19:45426792 | rs141622900 | A | 0.1007 |
| 19:46268902 | rs2341097 | T | 0.0502 |
| 19:49241006 | rs12981072 | C | 0.0469 |
| 19:55536595 | rs1613662 | A | 0.0818 |
| 19:7832001 | rs874492 | A | 0.0481 |
| 20:22672552 | rs6137727 | A | 0.0534 |
| 20:22938940 | rs62204096 | A | 0.1565 |
| 20:23000653 | rs6076004 | T | 0.0472 |
| 20:23077117 | rs149439892 | A | 0.1527 |
| 20:23170450 | rs34397775 | A | 0.084 |
| 20:23182559 | rs6083037 | A | 0.0798 |
| 20:32485961 | rs6087538 | C | 0.0585 |
| 20:32523172 | rs6059574 | G | 0.0479 |
| 20:33434252 | rs56244533 | A | 0.0567 |
| 20:33435161 | rs17092148 | G | 0.0612 |
| 20:33451060 | rs75627267 | T | 0.1209 |
| 20:33587569 | rs551986443 | C | 0.1488 |
| 20:33612647 | rs564783003 | C | 0.1488 |
| 20:33733180 | rs2050652 | G | 0.0564 |
| 20:33762035 | rs2069946 | C | 0.1255 |
| 20:33772243 | rs6060288 | A | 0.1178 |
| 20:33858772 | rs6058218 | G | 0.0552 |
| 20:33895947 | rs6058227 | T | 0.0813 |
| 20:34025983 | rs143383 | G | 0.0482 |
| 20:34562935 | rs185663249 | A | 0.1487 |
| 20:34712310 | rs6141600 | C | 0.0482 |
| 22:42461918 | rs2854827 | G | 0.0589 |
| 22:43107837 | rs9620086 | G | 0.0609 |
| 22:44324730 | rs738408 | C | 0.0566 |
| 1:169519049 | rs6025 | T | 0.4031 |
| 11:46761055 | rs1799963 | A | 0.2741 |

**sTable 2:**
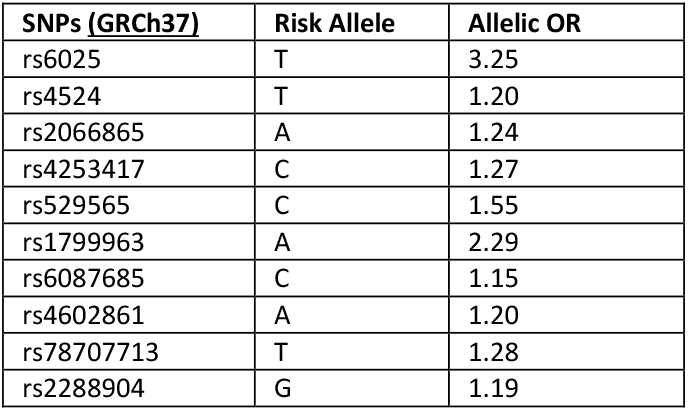
Summary statistics for candidate SNPs used for constructing sensitive polygenic risk score (Secondary PRS_VTE).

**sTable 3:**
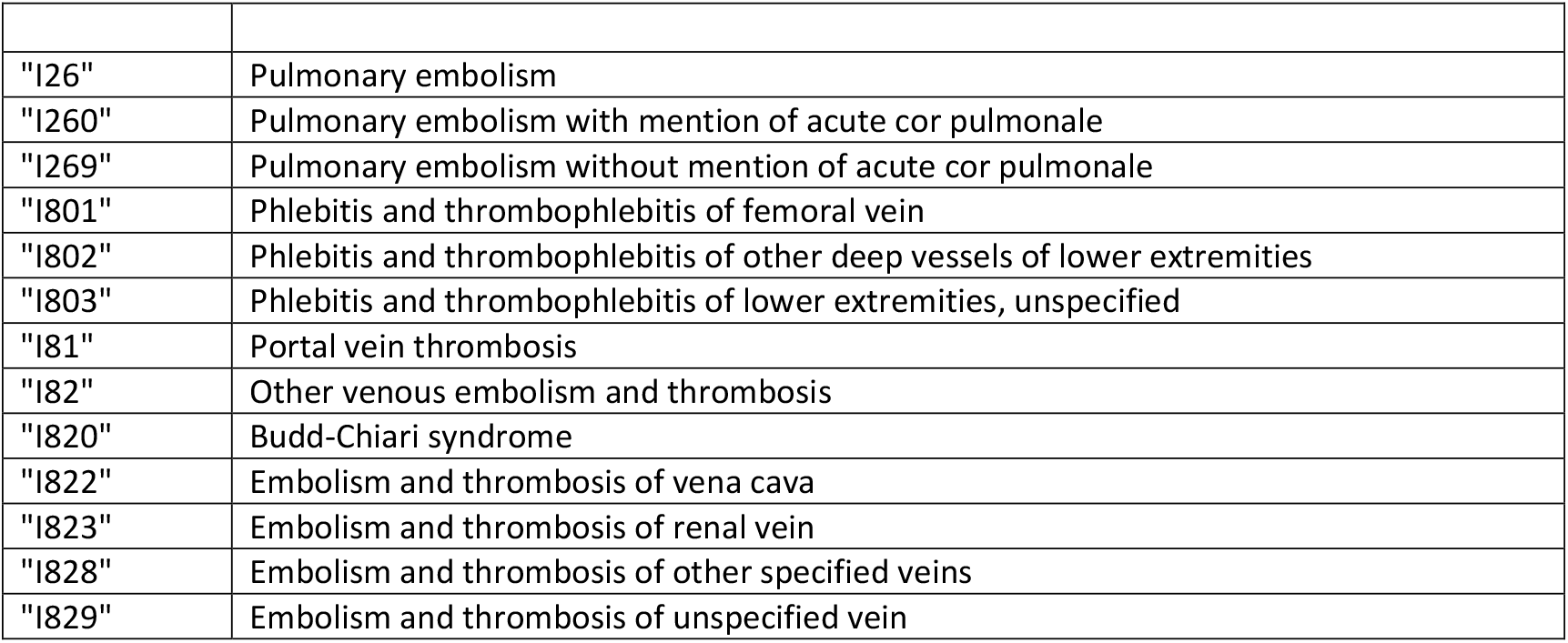
ICD-10 codes for the identification of venous thromboembolism.

**sTable 4:** Association between the polygenic risk score and the negative control outcome.

|  | Number<br>of people | Number<br>of cases | Incidence rate<br>(95% CI)* | Adjusted hazard<br>ratio (95% CI) <sup>§</sup> |
| --- | --- | --- | --- | --- |
| **Diabetes** |  |  |  |  |
| Whole UKBB (Pre-pandemic) | 373,207 | 2522 | 1.85 (1.78 to 1.93) | 1.02 (0.98 to 1.06) |
| Whole UKBB (Early-pandemic) | 368,112 | 1278 | 1.26 (1.20 to 1.34) | 0.98 (0.93 to 1.04) |

